# Brain-heart interactions in novice meditation practitioners during breath focus and an arithmetic task

**DOI:** 10.1101/2023.07.06.23292291

**Authors:** Javier R. Soriano, Julio Rodriguez-Larios, Carolina Varon, Nazareth Castellanos, Kaat Alaerts

**Author notes:** Corresponding Author: Prof. dr. Kaat Alaerts, KU Leuven, Neuromodulation Laboratory, Tervuursevest 101 - box 1501 3001 Leuven, Belgium.

## Abstract

**Objectives:** The study of neural and visceral oscillatory activities reveals that both subsystems and their interactions influence human cognition. In particular, cardiac and neural changes during self-regulation processes can be studied through a comparison of stress-inducing procedures and meditation practices.

**Methods:** In this study, we investigate the characteristic profiles of neural-cardiac interactions during a stress-inducing arithmetic task and a breath focus meditation period in a sample of 21 young participants (10 women, age range 20-29) with no prior experience in meditation practices. Using recordings of electroencephalography (EEG) and electrocardiography (ECG), we assessed instantaneous cross-frequency relationships between the alpha neural band and heart rate in both conditions.

**Results:** Our results indicate significant heart rate and alpha frequency decelerations during breath focus compared to the stress-inducing task. Regarding alpha: heart rate cross-frequency relationships, the stress-inducing arithmetic task exhibited ratios of smaller magnitude than the breath focus task, including a higher incidence of the specific 8:1 cross-frequency relationship, compared to the breath-focus task, proposed to enable cross-frequency coupling among neural and cardiac rhythms during mild cognitive stress. The change in cross-frequency relationships were mostly driven by changes in heart rate frequency between the two tasks, as indicated through surrogate data analyses.

**Conclusions:** Our results provide novel evidence that stress responses and changes during meditation practices can be better characterized by integrating physiological markers and, more crucially, their interactions. Together, this physiologically comprehensive approach can aid in guiding interventions such as physiology modulation protocols (biofeedback and neurofeedback) for emotion and stress-regulation.

## 1 INTRODUCTION

The overwhelming nature of contemporary life rhythms has fostered the proliferation of techniques aimed at enhancing self-regulation abilities. A predominant set of techniques employed to effectively cope with stressors can be found within meditation or contemplative practices (Chiesa & Serretti, 2009). Breath focus or mental scanning of body sensations, commonly taught in Mindfulness-Based Stress Reduction (MBSR) programs (Kabat-Zinn, 1990), are the most scientifically studied and practiced meditation techniques in western cultures. There is a vast body of research evidencing the benefits of meditation practices, which exert their effects through processes of attention regulation, body awareness, emotion regulation and shift in perspective of the self (Hölzel et al., 2011). Changes in brain function and structure as a result of contemplative practices have been widely investigated (Gotink et al., 2016; Hölzel et al., 2013; Tang et al., 2015). Furthermore, several studies have also shown mindfulness practices to result in down-regulation of stress reactivity (Goldin & Gross, 2010; Gotink et al., 2016; Kral et al., 2018), reductions in established physiological markers of stress including heart rate (Bortolla et al., 2022; Ooishi et al., 2021; Sun et al., 2019), reactivity of the autonomic nervous system and the stress hormone cortisol (Heckenberg et al., 2018), as well as reductions in other hormonal and immunological stress markers (Hoge et al., 2018). On the opposite end of the stress continuum, tasks such as mental arithmetic have been frequently used to study the physiological correlates of induced stress. Thus, characteristic profiles of neural activity have also been related to psychological stress (Lewis et al., 2007; Marshall et al., 2018; Marshall & Cooper, 2017; Zanetti et al., 2019). Other studies have also found heart rate (HR) to increase during mental arithmetic tasks (Sun et al., 2019; Watford et al., 2020), and the Trier Social Stress Test (TSST, Hoge et al., 2018).

Despite paving the way for unravelling the physiological mechanisms underpinning self-regulation through meditation practices, most research tends to focus on physiological subsystems independently (e.g. neural, cardiac). Although previously overlooked, science has recently started to reveal visceral influences on cognition and its neural underpinnings (Critchley & Garfinkel, 2018; Varga & Heck, 2017). For instance, studies have shown that cognitive experience is contingent to cardiac activity (Azevedo et al., 2017; Wilkinson et al., 2013) and its neural processing (Park et al., 2014). Interestingly, regarding cardiac influences on neural activity during meditation practices, Jiang and colleagues (2020) observed transient modulations of the neural response to heartbeats in the default mode network (DMN), as well as a stronger directional coupling from the gamma to the theta band. In relation to other visceral influences, different breathing characteristics such as frequency or modality (oral or nasal) have also been shown to affect neural oscillations and cognitive processing (Arshamian et al., 2018; Heck et al., 2019; Herrero et al., 2018; Hsu et al., 2020; Lee et al., 2020; Perl et al., 2019; Zelano et al., 2016). Thus, to address the inherent systemic nature of self-regulation, and to acknowledge the complexity arising from dynamic interactions between human physiological subsystems, the approach to study physiological processes is shifting towards a systemic domain, utilizing multivariate and network analysis methods (Bartsch et al., 2015; Bashan et al., 2012). Several methodologies are therefore being developed to analyze simultaneous physiological signals, applying tools deriving from fields such as Information dynamics, Bayesian inference or synthetic data generation (Bartsch et al., 2015; Bashan et al., 2012; Candia-rivera et al., 2023; Candia-Rivera, Catrambone, Barbieri, et al., 2022; Candia-Rivera, Catrambone, Thayer, et al., 2022; Stankovski et al., 2016; Zanetti et al., 2019), as well as neural event related potentials such as the heartbeat evoked potential (HEP) (Jiang et al., 2020; Park & Blanke, 2019).

Physiological activity is commonly studied through the perspective of its oscillatory or rhythmic patterns. In the case of brain activity, electroencephalography (EEG) allows to observe oscillations arranged in distinct frequency bands. As elements of the same physiological subsystem, these oscillations inextricably interact to enable communication within the brain across different spatio-temporal scales, hence allowing for complex information processing (Canolty & Knight, 2010; Fries, 2015; Hyafil et al., 2015; Varela et al., 2001). The Binary Hierarchy Brain Body Oscillation Theory (Klimesch, 2018), follows up on this neurally observed interactions, and presents a model of a unified architecture comprised by the main oscillations, both neural and visceral, in the human body. This theory proposes that different body subsystem oscillations interact via the same oscillatory coupling principles of cross-frequency phase coupling, allowing communication across different spatio-temporal scales. The ability of two oscillators to phase couple depends on whether their respective frequencies maintain a harmonic/integer cross-frequency relationship. For example, at any given time and within their frequency ranges, the neural oscillations alpha (α) and theta (θ) can oscillate at 12 and 6 Hz respectively, yielding a harmonic relationship of 2:1. In a state and task dependent manner, and specifically upon cognitively demanding conditions, neural oscillations have been shown to increase phase coupling (Palva et al., 2005; Rodriguez-Larios & Alaerts, 2019; Sauseng et al., 2008; Siebenhühner et al., 2016). In the context of MBSR interventions and meditation practices, the analyses to test cross-frequency relationships between alpha and theta rhythms have been further developed and employed in a series of studies (Rodriguez-Larios, Faber, et al., 2020; Rodriguez-Larios, Wong, et al., 2020; Rodriguez-Larios & Alaerts, 2019, 2020). These studies have revealed that the occurrence of harmonic frequency relationships and phase coupling between alpha and theta is higher during a stress-induction arithmetic task compared to rest and breath focus periods (Rodriguez-Larios & Alaerts, 2019). This latter study and subsequent ones (Rodriguez-Larios, Faber, et al., 2020; Rodriguez-Larios, Wong, et al., 2020; Rodriguez-Larios & Alaerts, 2020), also show that harmonic cross-frequency relationships and phase coupling are less pronounced during meditation practices when compared to rest and an arithmetic task in expert meditators (Rodriguez-Larios, Faber, et al., 2020). Similarly, the degree of attendance to an MBSR program has been shown to predict decreases in harmonicity and phase coupling between alpha and theta oscillations during meditation practices (Rodriguez-Larios, Wong, et al., 2020). Even in novice meditators, it is shown that during a period of breath focus meditation, self-reported mind-wandering (distracting self-generated thoughts) is associated with an increase in harmonic cross-frequency relationships and phase coupling between alpha and theta compared to periods of breath focus (Rodriguez-Larios & Alaerts, 2020). These studies suggest that the non-harmonicity relationships characteristic of meditation practices preclude the unwanted interaction between executive and memory processes, aiding the achievement of a meditative state (Rodriguez-Larios, Faber, et al., 2020).

Cross-frequency dynamics among neural and visceral rhythms (i.e., alpha rhythm and heart rate/respiration) however, have only been investigated in one study (Rassi et al., 2019). Considering the aforementioned effects of visceral physiology in cognition and neural activity, and with the aim of testing Klimesch’s (2018) theory, Rassi and colleagues (2019) studied cross-frequency ratios between alpha and heart rate, alpha and respiration, as well as heart rate and respiration during a memory task, rest and sleep. Cross-frequency ratios derived from the average peak frequencies of the oscillators, revealed a higher occurrence of harmonic frequency relationships between alpha, heart rate and breathing frequency during a memory task, but a non-harmonic neural-visceral relationship during rest and sleep.

In the current study we assess the relevance of the Binary Hierarchy Brain Body Oscillation Theory (Klimesch, 2018; Rassi et al., 2019) through an examination of changes in neural-cardiac cross-frequency arrangements during breath focus practice versus a stress-inducing arithmetic task in novice meditators. Crucially, our study approaches the assessment of cross-frequency arrangements from an instantaneous, instead of an average perspective (Rassi et al., 2019). Thus, we developed a novel examination of transient changes in the alpha and heart rate rhythms as well as their cross-frequency ratio, allowing a fine-grained analysis of time-varying dynamics in harmonic versus non-harmonic cross-frequency arrangements.

## 2 METHODS

### 2.1 Participants

Twenty-eight healthy volunteers (11 males, mean age 23.46 years, age range: 20–29) were recruited to participate in the study. Informed written consent was obtained from all participants before the study. Consent forms and study design were approved by the Social and Societal Ethics Committee (SMEC) of the KU Leuven university (G-2018 12 1,463), in accordance with the World Medical Association Declaration of Helsinki. Participants were compensated for their participation (8€ per hour). Participants took part in all conditions of the study, except for two participants who did not complete one of the two conditions of interest. Five additional participants were excluded due to technical problems during data acquisition, and another participant due to low data quality, rendering the total number of analysed participants to 20 (12 females, average age: 23,38, age range: 20-29).

Note that participants of the current study were recruited to participate in a larger project additionally including a rest condition and a heartbeat counting task, as well as a probe-caught mind wandering focus task, previously described in Rodriguez-Larios & Alaerts (2020).

### 2.2 Design and tasks

Continuous electroencephalographic (EEG) and electrocardiographic (ECG) recordings were simultaneously obtained while participants performed an unguided breath focus session and a stress-inducing arithmetic task. During the breath focus session, similar to instructions given in MBSR programs (Kabat-Zinn, 1990), participants were instructed to focus on the sensation of breathing, for approximately 5 minutes, while closing their eyes, and to come back to that focus whenever they noticed their mind wandered off in thoughts. The arithmetic task condition lasted approximately 5 minutes, and was divided in four trials. In each trial, participants were instructed to reproduce a numerical series similar to Fibonacci’s in their minds. Thus, they were required to iteratively add two numbers, and subsequently add the result to the second term of the previous addition (i.e., starting with numbers 0 and 1; 0 +1 = 1, 1 + 1 = 2, 1 + 2 = 3, 2 + 3 = 5…), until their mental calculation surpassed 100 and, once reached, they were required to press a button and verbally inform the experimenter about the obtained result. Accordingly, the total length of the arithmetic task varied depending on how fast each participant performed the calculations and responded in each trial.

### 2.3 Data acquisition and analysis

We assessed the relationship between neural and cardiac electrophysiology during conditions of breath focus and the stress-inducing mental arithmetic task. To that end, we employed an alpha frequency band (8-14) and heart rate frequency (0.6-2 Hz) instantaneous cross-frequency ratio approach.

#### 2.3.1 Recordings

The Nexus-32 system (version 2015a, Mind Media, The Netherlands) and BioTrace software (Mind Media) were used to record EEG and ECG.

*EEG.* Continuous EEG was recorded with a 22-electrode cap (two reference electrodes and one ground electrode) positioned according to the 10–20 system (MediFactory). Vertical (VEOG) and horizontal (HEOG) eye movements were recorded by placing pre-gelled foam electrodes (Kendall, Germany) above and below the left eye (VEOG) and next to the left and right eye (HEOG) (sampling rate of 2048 Hz). Skin abrasion and electrode paste (Nuprep) were used to reduce the electrode impedances during the recordings. The EEG signal was amplified using a unipolar amplifier with a sampling rate of 512 Hz.

*ECG.* Continuous ECG was recorded with a bipolar two-lead electrode configuration. Active electrodes were placed below the left ribcage and below the right collarbone and ground electrode was that of the EEG cap. Signal was amplified using a bipolar amplifier with a sampling rate of 256 Hz. Recordings were synchronized to the presented tasks using E-prime and the Nexus trigger interface (Mind Media).

#### 2.3.2 Pre-processing

EEG pre-processing was performed through custom MATLAB scripts and EEGlab functions (Delorme & Makeig, 2004) and was similar to that described in Rodriguez-Larios & Alaerts (2020). First, the initial and final two seconds of raw EEG data were rejected, and remaining data was down sampled to 256 Hz to match the sampling rate of ECG data. Then a bandpass filter between 0.5 Hz and 40 Hz was used to attenuate non-physiological EEG artifacts (function *pop_eegfiltnew*). Noisy channels were automatically detected based on the correlation with their robust estimate (see function *clean_channels;* correlation threshold = 0.6). EEG was re-referenced to common average and independent component analysis (ICA) was performed to reject activity related to eye movements, muscle activity and cardiac artifacts (*runica* algorithm as implemented in EEGlab; number of components was equal to the number of non-interpolated channels minus 1 to avoid rank deficiency). Components to be rejected were automatically detected using the *iclabel* classifier (Pion-Tonachini et al., 2019). EEG data was divided into 1-s epochs and those exceeding an absolute amplitude of 50 μV were rejected.

To avoid signal to noise ratio differences between the two experimental conditions (breath focus and arithmetic), the number of epochs was matched in subsequent analyses by selecting the initial epochs of the condition with a higher number of clean epochs to match the number of the condition with a lower number of clean epochs (see Table 1 in Supplementary Material).

ECG was pre-processed by first rejecting the initial and final two seconds of raw data, then data was zero-phase filtered between 1 and 20 Hz (Butterworth filter with order 2 and 4 respectively) using the Matlab function *filtfilt*. R peaks within the QRS complex were automatically detected and annotated with the Matlab toolbox R-DECO (Moeyersons et al., 2019). When needed, automatic R peak annotations were corrected by visual inspection.

#### 2.3.3 | Condition-related differences in average and transient alpha: heart rate cross-frequency ratios

An instantaneous cross-frequency ratio approach was employed (similar to Rodriguez-Larios et al., 2020) for the calculation and comparison of frequency relationships between alpha and heart rate during breath focus and arithmetic task conditions.

*EEG transient alpha peak frequency.* First, short-term fast Fourier transformations were performed in each 1 second EEG epoch using the function *spectrogram* in MATLAB 2019b (0.1 Hz resolution between 0.1 and 40 Hz). Thereafter, transient peak frequencies in the alpha frequency band (8-14 Hz) were extracted per second using the MATLAB function *findpeaks*, which define peaks as data samples that are larger than its two neighbouring samples. When more than one peak was detected, the peak with the highest amplitude was selected. Thus, per condition, participant and electrode, one alpha peak frequency time series was calculated.

*ECG transient heart rate frequency.* From the annotated R peaks, inter beat intervals (IBIs) were calculated and used to derive instantaneous heart rate in Hz, dividing IBIs by 60.

*Transient Alpha: heart rate cross-frequency ratio (see Figure 1)*. Dividing the alpha frequency time series by the heart rate frequency time series, the cross-frequency ratio series were calculated per condition, participant, and electrode. With an a priori definition of alpha between 8-14 Hz and heart rate typically oscillating between 0.6 - 2 Hz, computed ratios ranged between 4 and 24, with a resolution of 0.5 (41 possible ratios between the two oscillators). Thereafter, the percentage of occurrence of each possible ratio within the ratio time series was calculated per condition, electrode, and participant.

**Figure 1.**
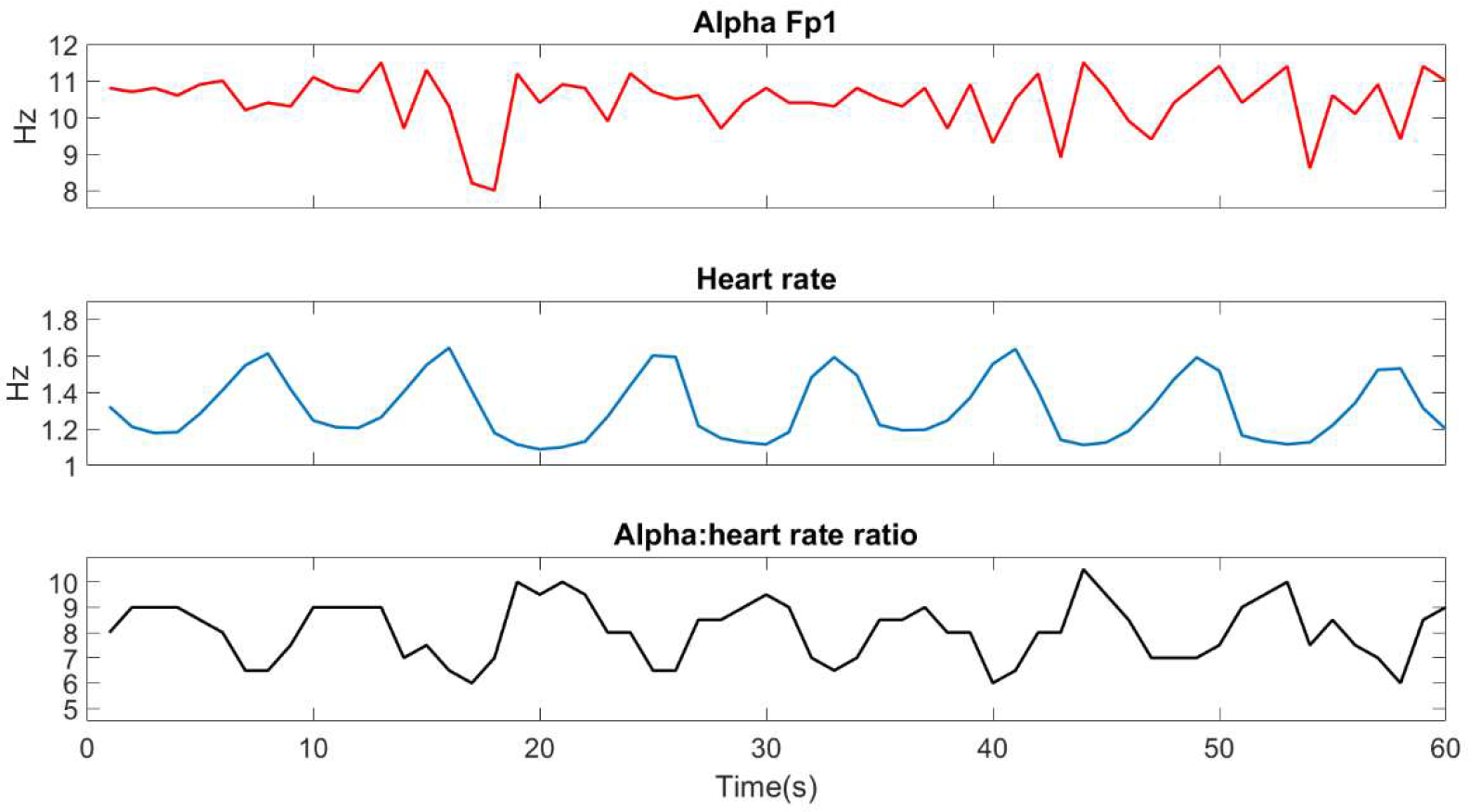
Electrophysiological one-minute recording of an exemplary participant. Visualization of neural alpha (top panel, example electrode Fp1) and heart rate (middle panel) instantaneous frequency time series, from which the instantaneous cross-frequency relationship (bottom panel, ratio alpha: heart rate) is calculated.

In addition to the instantaneous approach, also ‘average’ cross-frequency relationships were obtained (similar to Rassi et al., 2019), by calculating the average individual alpha frequency (IAF), by averaging the previously obtained spectrograms across time, and finding a peak comprehended between 8 and 14 Hz, as well as the average heart rate. Next, the average cross-frequency ratio between the average alpha and heart rate frequencies was calculated for each condition, electrode, and participant.

#### 2.3.4 Condition-related differences in alpha: heart rate cross-frequency ratios with surrogate data

In order to control for differences between conditions in the two independent physiological signals (alpha frequency and heart rate) used to compute cross-frequency ratios, as well as for the relevance of physiologically recorded instantaneous alpha: heart rate relationships, four surrogate data approaches were used.

*Surrogate 1, permutated heart rate across conditions.* First, a distribution of recorded heart rate frequencies per participant was derived combining values from the time series of both conditions (breath focus and arithmetic). Then, the moments (mean, standard deviation, kurtosis and skewness) of the combined distribution were calculated and used to create surrogate heart rate time series per participant, using the function *pearsonrnd*, which draws values from a distribution with the specified moments. While maintaining the original alpha frequency time series per participant, new cross-frequency ratios and their percentage of occurrence were calculated using the surrogate heart rate time series.

*Surrogate 2, permutated alpha frequency across conditions.* The same procedure was used to calculate surrogate alpha frequency time series per participant and electrode, while maintaining original heart rate time series. Then, new cross-frequency ratios and their percentage of occurrence were calculated.

Permutating either heart rate (surrogate 1) or alpha frequency time series (surrogate 2) across the breath focus and stress-induction conditions, maintaining the same distribution moments across conditions in one of the signals, allowed examining the relative contribution of each physiological signal in driving the identified condition-related differences in alpha: heart rate cross-frequency relationships. If condition-related differences in cross-frequency occurrences do not prevail after permutating the alpha frequency or heart rate time series across conditions, it can be inferred that condition-related differences in either single time series were necessary to yield condition-specific effects in the cross-frequency arrangements. Conversely, if condition-related differences persist with the surrogate time series, it can be inferred that changes in the maintained original physiological signal were enough in driving the condition-related effects in cross-frequency ratios.

Next, similar surrogate analyses were performed, permutating either alpha or heart rate to examine their respective contribution to yielding observed patterns of condition-related effects in cross-frequency occurrence, albeit now, random heart rate (surrogate 3) or alpha frequencies (surrogate 4) were calculated within the defined frequency ranges. Thus, omitting physiologically relevant characteristics of the distribution of observed alpha or heart rate frequencies. These surrogate analyses thereby allow examining the relative contribution of physiologically observed alpha frequency or heart rate distributions in yielding the condition-related differences in alpha: heart rate cross-frequency ratios.

*Surrogate 3, random heart rate*. In this surrogate approach, random values comprehended between 0.6 and 2 were drawn, to produce random heart rate series per participant, while maintaining original alpha frequency. Such series was then used to compute new cross-frequency ratios and their percentage of occurrence, using original alpha frequency time series per participant.

*Surrogate 4, random alpha*. The same procedure was used selecting random values between 8 and 14 to produce random alpha frequency series per participant and electrode, while maintaining original heart rate series to compute cross-frequency ratios and associated percentages.

For each of the four surrogate controls, the procedure was repeated 1000 times and used for further analyses (see Section Statistical Analyses). Results for the surrogates 3 and 4 are shown in Supplementary Materials.

#### 2.3.6 Statistical analyses

*Average condition differences*. To compare the average values of heart rate, IAF and IAF: heart rate cross-frequency ratio between conditions, we applied paired-sample t-test with the function ttest in MATLAB.

*Cluster-based permutation tests.* For each of the computed variables of interest (ratio percentages) a cluster-based permutation statistical method was adopted to assess condition-related differences (between breath focus and arithmetic task) as implemented in the MATLAB toolbox Fieldtrip (function ft_timelockstatistics) (Maris & Oostenveld, 2007). This statistical method controls for the type I error rate arising from multiple comparisons through a non-parametric Montecarlo randomization. In short, data were shuffled (10,000 permutations) to estimate a ‘null’ distribution of effect sizes based on cluster-level statistics (sum of t-values with the same sign across adjacent electrodes and ratios). Then, the cluster-corrected p-value was defined as the proportion of random partitions in the null distribution for which the test statistics exceeded the one obtained for each significant cluster (cluster-defining threshold: p < .05) in the original (non-shuffled) data. Significance level for the cluster permutation test was set to 0.025 (corresponding to a false alarm rate of 0.05 in a two-sided test) (Maris & Oostenveld, 2007). To assess condition-related differences, paired-sample t-test (function ft_statfun_depsamplesT in Fieldtrip) was chosen as the test statistic.

*Surrogate data analyses*. For each of the four surrogate data analyses and their respective 1000 repetitions, the same cluster-based permutation approach as with original data was employed to test for significance. A distribution of average t-values, as well as number of times that each cell (electrode by ratio) came significant within a cluster was calculated. Next, to test the significance of t-values from the permutation test with original data, against the surrogate distribution of 1000 t-values, the normal cumulative density function (Matlab function *normcdf*) of the 1000 values was calculated. Each of the original t-values within the electrode by ratio grid, was then searched within the surrogate distribution to select the associated p-value. The original t-value was marked as significant if its associated p-value in the cdf was below 0.025 or above 0.975. Subsequently, to account for the multiple comparisons, the grid of associated p-values was FDR (false discovery rate) corrected (following the procedure of Benjamini and Yekutieli, 2001) with threshold *q* set at 0.05.

Additionally, relationships between condition-related changes from breath focus to arithmetic task in the individual physiological measures (heart rate, alpha and IAF) and the percentages across relevant ratios were examined via Pearson correlations, calculated using the *corrplot* function in Matlab.

## 3 RESULTS

### 3.1 Condition-related differences in alpha and heart rate electrophysiological measures

First, we explored condition-related modulations in each of the physiological measures separately. As depicted in **Figure 2A**, most participants showed a significantly lower average heart rate in the breath focus condition compared to the stress-inducing arithmetic condition. Significance of this difference was evaluated using a paired-sample t-test (*t*(19) = 5.22; *p* < 0.01) on the participant average values per condition (mean breath focus = 1.10 Hz; *SD* = 0.13; mean arithmetic = 1.20 Hz; *SD* = 0.16). Condition-related differences in individual alpha frequency (IAF) were also present (**Figure 2B**), indicating lower IAF in the breath focus condition compared to the arithmetic condition. As visualized in the topographical plot in **Figure 2B**, marked with asterisks, the condition-related difference in IAF was particularly evident at electrodes clustered towards right-and central-frontal locations (Fz, F4, Cz) (t_cluster_ (19) = 9.14; p = 0.01).

**Figure 2.**
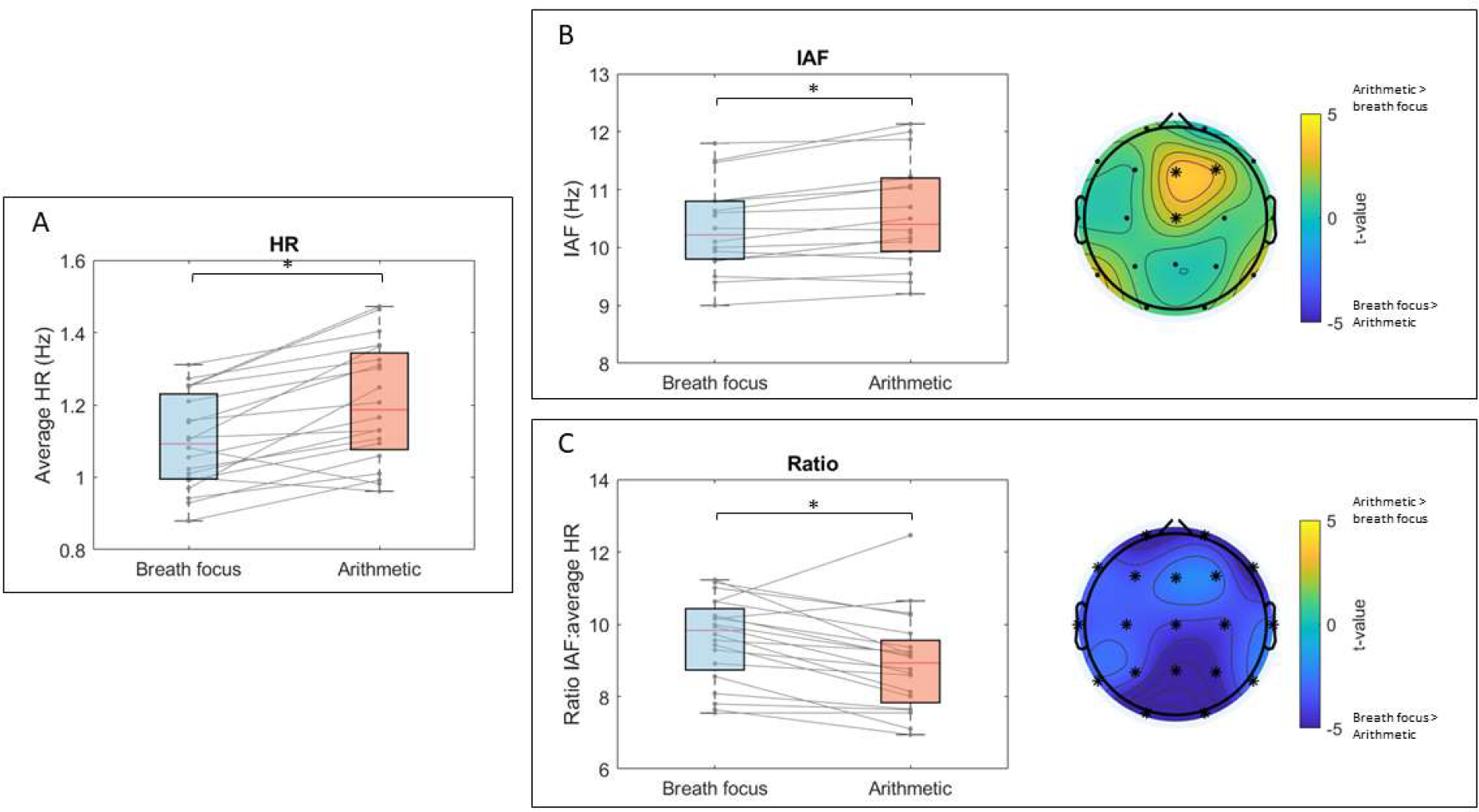
Condition-related differences in electrophysiological measures. For each measure and condition, box plots are presented, depicting median (red line), minimum and maximum values (bottom and top whiskers) as well as first (bottom) and third quartiles (top box boundary). **A)** visualizes average heart rate (HR), separately for the breath focus (blue) and arithmetic conditions (red). **B)** visualizes average individual alpha frequency (IAF) (average across significant electrodes), separately for the breath focus (blue) and arithmetic conditions (red). The adjacent topographical plot shows the t-values of condition-related differences (arithmetic-breath focus), as well as electrodes showing significant differences in IAF between conditions (*p<0.05). **C)** visualizes cross-frequency ratio averages (IAF: average HR) (across significant electrodes) separately for the breath focus (blue) and arithmetic conditions (red). Also here, the adjacent topographical plot depicts the t-values of condition-related differences, with indication of significant electrodes with an asterisk (*p<0.05).

### 3.2 Condition-related differences in average alpha: heart rate cross-frequency ratios

Average cross-frequency ratios (IAF: average heart rate), calculated for each electrode, were shown to be significantly higher for the breath focus condition (mean = 9.58; SD = 1.16) compared to the arithmetic condition (mean = 8.92; SD = 1.34) (t_cluster_(19) = −78.89 ; p < 0.001). As visualized in **Figure 2C**, the effect was evident for all electrodes (marked with asterisks).

### 3.3 Condition-related differences in transient alpha: heart rate cross-frequency ratios

Next, we explored whether condition-related modulations in instantaneous cross-frequency ratios were present. In **Figure 3**, distributions of average percentage (across participants) of each possible ratio at each electrode location are depicted. During the breath focus condition (**Figure 3A**), cross-frequency ratios followed a normal distribution, similar across the entire scalp, with a maximal percentage at ratio 9 (11.11%, averaged across electrodes). Also, during the arithmetic condition, cross-frequency ratios followed a normal distribution with maximal occurrence at ratio 8.5 (13.42%) (**Figure 3B**). Cluster-based permutation statistics examining condition-related differences (**Figure 3C**) revealed two significant clusters. One cluster (grouping positive t-values, arithmetic > breath focus) indicated that - across the entire scalp - the occurrence of ratios between 6 and 8.5 was significantly higher during the arithmetic condition (Mean _cluster_ = 8.26%; SD = 4.59) compared to the breath focus condition (Mean _cluster_ = 5.24%; SD = 3.4) (t _cluster_ (19) = 183.9 ; *p* = 0.004). Interestingly, this cluster includes the harmonic alpha: heart rate cross-frequency ratio of 8, as hypothesized by Klimesch (2018). Conversely, another cluster (grouping negative t-values, arithmetic < breath focus) indicated that the occurrence of ratios between 10 and 13.5 was significantly higher during the breath focus condition (Mean _cluster_ = 5.48%; SD = 3.36) compared to the arithmetic condition (Mean _cluster_ = 3.33%; SD = 2.77), also across all electrodes (t _cluster_ (19) = −286.82 ; *p* < 0.001). Also in this case, the hypothesized non-harmonic ratio of 12.94 is within the ratio range of the cluster. Note that, due to the absence of data for ratios smaller than 4.5 and greater than 18.5 in both conditions (**Figure 3A and 3B**), the permutation analysis was performed on a subset of the ratios (4-19) leaving a relative margin in both data boundaries (**Figure 3B**).

**Figure 3.**
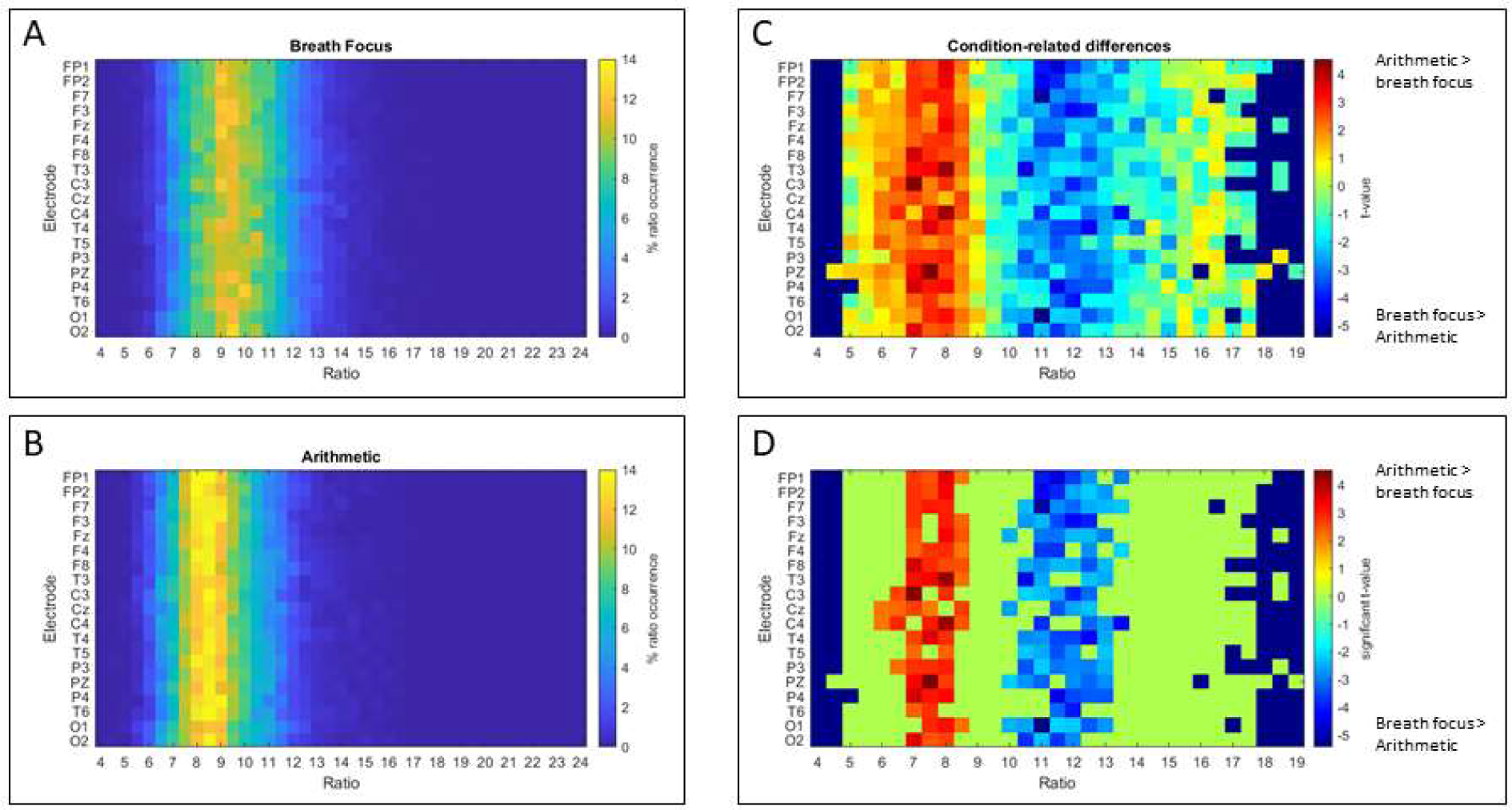
Condition-related differences in the incidence (% of epochs) of different alpha: heart rate cross-frequency ratios. **A)** visualizes the distribution of the percentage of each ratio occurrence for the breath focus condition, separately for each electrode (y-axis) and cross-frequency ratio (x-axis) (ratios ranging between 4 and 24). **B)** visualizes the distribution of the percentage of each ratio occurrence for the arithmetic condition. **C)** visualizes the colormap of t-values derived from cluster-based permutation analysis estimating the condition-related effect (paired-sample t-test; arithmetic versus breath focus condition), separately for each electrode (y-axis) and cross-frequency ratio (x-axis). Permutation analyses were performed for ratios ranging between 4 and 19, since the occurrence of ratios below 4.5 or above 18.5 was < 0.01 %. **D)** shows the map of t-values masked for significance after the multiple comparison correction of the cluster-based permutation (*p<0.025).

### 3.3 Condition-related differences in alpha: heart rate cross-frequency ratios with surrogate data

Given the significant difference in heart rate across conditions, and to assess the robustness of the instantaneous alpha: heart rate ratio approach compared to the average IAF: heart rate approach (**Figure 2**), a surrogate data procedure was employed. Specifically, four surrogate analyses were performed, (i) one in which alpha: heart rate ratios were calculated with surrogate heart rate data derived from a distribution with the same moments as that of both conditions combined, and original alpha data (surrogate 1 heart rate, **Figure 4**); (ii) the other in which alpha: heart rate ratios were calculated with surrogate alpha data derived from a distribution with the same moments as that of both conditions combined, and original heart rate data (surrogate 2 alpha, **Figure 5**). (iii) one in which alpha: heart rate ratios were calculated with surrogate heart rate data randomly assigned between 0.6 and 2 Hz, and original alpha data (surrogate 3 heart rate, **Supplementary Figure 1**); (iv) the other in which alpha: heart rate ratios were calculated with surrogate alpha data randomly assigned between 8 and 14 Hz, and original heart rate data (surrogate 4 alpha, **Supplementary Figure 2**).

**Figure 4.**
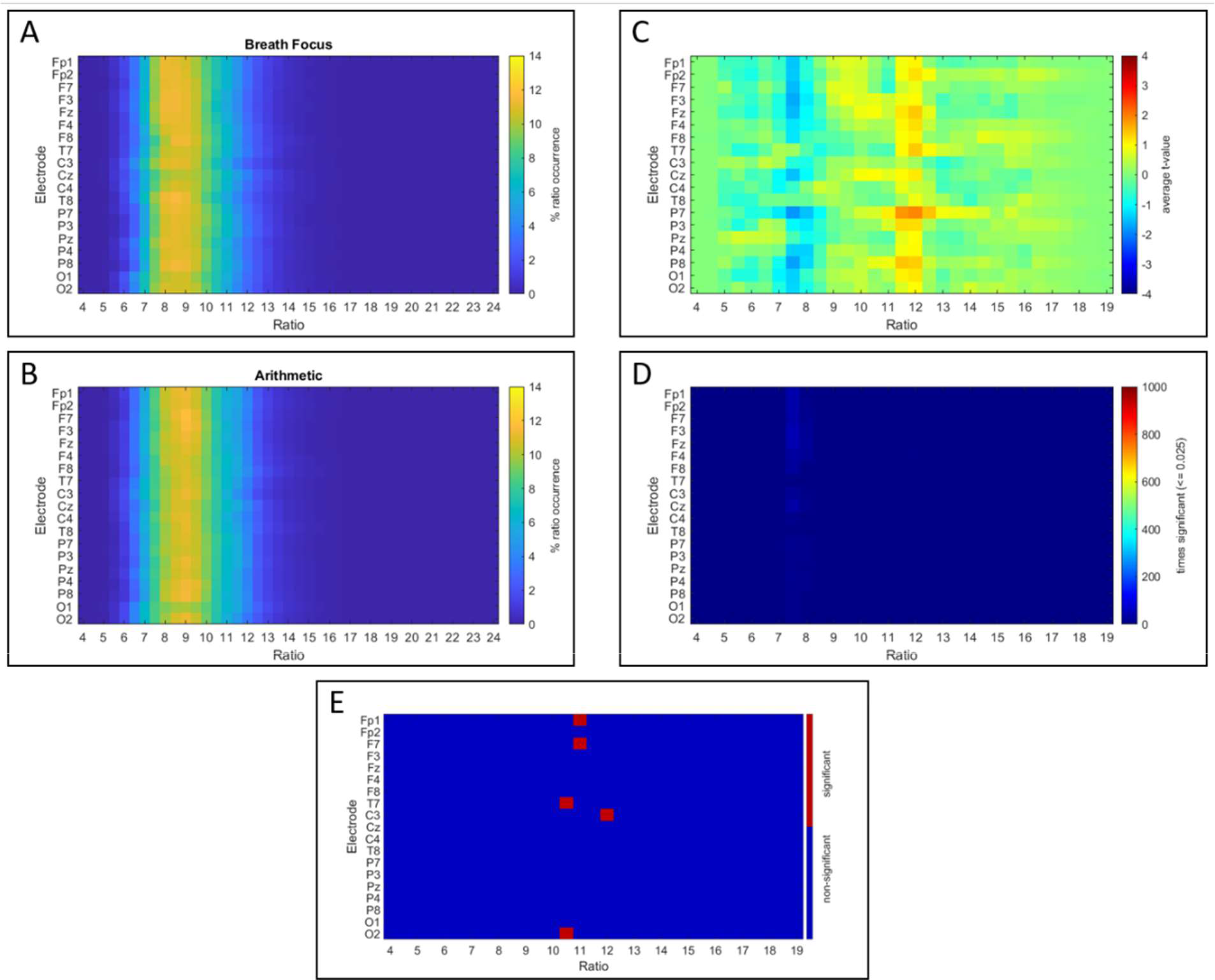
Condition-related differences in alpha: heart rate cross-frequency ratios with heart rate surrogate data. **A)** visualizes the average distribution, across 1000 surrogate generations, of the percentage of each ratio occurrence for the breath focus condition, separately for each electrode (y-axis) and cross-frequency ratio (x-axis) (ratios ranging between 4 and 24). **B)** visualizes the distribution of the percentage of each ratio occurrence for the arithmetic condition. **C)** visualizes the colormap of the average t-values across the 1000 surrogate calculations and permutation tests estimating the condition-related effect (arithmetic versus breath focus condition), separately for each electrode (y-axis) and surrogate cross-frequency ratio (x-axis). Permutation analyses are visualized for ratios ranging between 4 and 19, since the occurrence of ratios below 4.5 or above 18.5 was < 0.01 %. **D)** visualizes the accumulated times that each of the cells (electrode-ratio t-test value) was significant, i.e., over the 1000 surrogate data generations, **E)** shows the significance of the original t-values (plotted in Figure 2D) against the t-value distribution derived from the 1000 surrogate generations.

**Figure 5.**
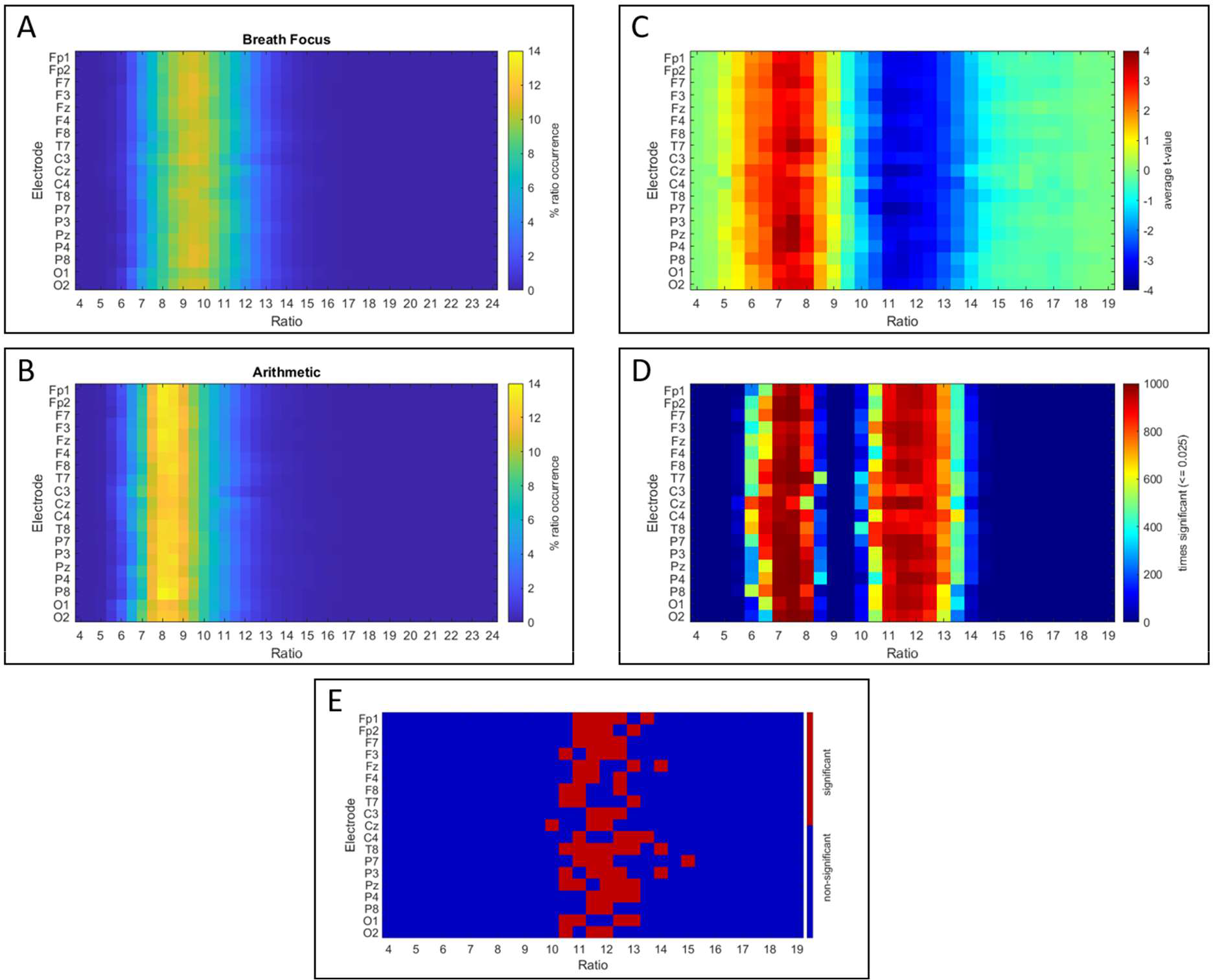
Condition-related differences in alpha: heart rate cross-frequency ratios with alpha surrogate data. **A)** visualizes the average distribution, across 1000 surrogate generations, of each ratio occurrence for the breath focus condition, separately for each electrode (y-axis) and cross-frequency ratio (x-axis) (ratios ranging between 4 and 24). **B)** visualizes the distribution of the percentage of each ratio occurrence for the arithmetic condition. Note that the percentage of occurrence of each ratio was calculated separately per participant and electrode, and subsequently averaged across participants. **C)** visualizes the colormap of the average t-values across the 1000 surrogate calculations and permutation tests estimating the condition-related effect (arithmetic versus breath focus condition), separately for each electrode (y-axis) and surrogate cross-frequency ratio (x-axis). Permutation analyses are visualized for ratios ranging between 4 and 19, since the occurrence of ratios below 4.5 or above 18.5 was < 0.01 %. **D)** visualizes the accumulated times that each of the cells (electrode-ratio t-test value) was significant, i.e., over the 1000 surrogate data generations, **E)** shows the significance of the original t-values (plotted in Figure 2D) against the t-value distribution derived from the 1000 surrogate generations.

*Surrogate 1, permutated heart rate across conditions.* In **Figure 4**, the average distribution of the 1000 surrogate calculations are shown for the breath focus and arithmetic conditions. During the breath focus condition (**Figure 4A**), cross-frequency ratios followed a normal distribution, similar across the entire scalp, with a maximal percentage at ratio 8.5 (11.12%, averaged across electrodes). Also, during the arithmetic condition, cross-frequency ratios followed a normal distribution with maximal occurrence at ratio 9 (11.1%) (**Figure 4B**).

Based on the heart rate surrogate data, only 61 times out of the 1000 repetitions (**Figure 4 C and D**), a significant negative cluster was identified – overlapping with the positive cluster identified in the original analysis –, indicating a higher occurrence of cross-frequency ratios, comprehended between 7 and 8.5, in the breath focus, compared to the arithmetic condition. Only in 12 times out of the 1000 surrogates, a positive cluster overlapping with the negative cluster of the original analysis was found, indicating a higher occurrence of ratios between 9.5 and 13 in the arithmetic, compared to the breath focus condition. This surrogate analysis therefore demonstrates that randomizing the time series of the heart rate data poses a significant impact on the identified condition-related differences in alpha: heart rate cross-frequency ratios between the breath focus and arithmetic condition, abolishing any condition-related effects.

A direct exploration, comparing the original permutation test t-value grid (presented in Figure 3D) with the surrogate 1000 t-value distribution, confirms the robustness of specific elements in the identified clusters of the original analysis, compared to the surrogate 1000 t-value distribution (see Statistical Analyses section). Particularly, after FDR correction (**Figure 4E**), t-values pertaining to ratio 10.5 in electrodes T7 and 02, ratio 11 in Fp1 and F7 and ratio 12 in C3 remained significant. This analysis therefore confirms that an intact heart rate time series forms a necessary prerequisite for yielding the identified significant condition-related effects in alpha: heart rate cross-frequency ratios.

*Surrogate 2 permutated alpha across conditions.* In **Figure 5A and B**, the average distribution of the 1000 surrogate calculations are shown for the breath focus condition and the arithmetic condition. During the breath focus condition (**Figure 5A**), cross-frequency ratios followed a normal distribution, similar across the entire scalp, with a maximal percentage at ratio 9.5 (10.71% averaged across electrodes). During the arithmetic condition, cross-frequency ratios followed a normal distribution with maximal occurrence at ratio 8 (12.77%) (**Figure 5B**).

Across the 1000 surrogates, a significant positive cluster (*p*<.025) was found 1000 times, with boundaries comprehended between the ratios 5 and 9, and typically spanning the entire scalp (**Figure 5D**). Negative clusters were also found 1000 times, between the ratios 9.5 and 15 across the entire scalp. Next, we tested the original permutation test t-value grid with the surrogate 1000 t-value distribution. After FDR correction, only t-values pertaining to the negative cluster (i.e. ratios significantly more frequent during breath focus compared to arithmetic) between ratios of 10 and 15 remain significant, indicating the robustness of that condition-related difference when tested against a condition of alpha surrogate data altering the physiological instantaneous relationships between heart rate and alpha frequency as well as abolishing the condition differences in alpha frequency. **Figure 5E** shows the two-tailed significance (*p*<.025) of the t-values, FDR corrected for multiple comparisons.

Together, these surrogate analyses demonstrate that permutating originally measured heart rate time series across conditions, but not alpha frequencies, abolishes identified condition-related effects in alpha: heart rate cross-frequency arrangements. This indicates that the condition-related difference in alpha: heart rate ratios rely heavily on instantaneous condition-specific changes in heart rate, and to a lesser extent on instantaneous condition-specific changes in alpha.

Omitting the physiologically relevant characteristics of the distribution of heart rates (surrogate 3, see supplement Figure 1), by randomly selecting heart rates between 0.6-2 Hz but with intact alpha frequency time series, was also sufficient to abolish the identified condition-related effects in alpha: heart rate cross-frequency arrangements. However, when generating random alpha frequencies between 8-14 Hz (surrogate 4, see supplement Figure 2), both positive and negative clusters were significant 100% of the permutations. Due to the difference in variability in the ratio distributions, with higher spread for the breath focus condition, the clusters that turn significant for the breath focus distribution, vary across permutations, whereas for the arithmetic task, the same turn significant every permutation (indicated in dark red). This indicates that, while condition-specific *instantaneous physiological* changes in alpha might contribute little to the observed condition-related changes in the original alpha: heart rate ratios (as seen in Figure 4), the overall distribution of physiologically recorded alpha appears to carry relevant information.

Accordingly, the participant-specific features of the physiologically alpha peak frequency distributions are likely relevant in relation to the participant-specific heart rate distributions for yielding the observed condition-related changes in alpha: heart rate cross-frequency arrangements.

### 3.5 Relationships between condition-related changes from breath focus to arithmetic task

Changes in alpha peak frequency or IAF from breath focus to arithmetic did not correlate with changes in heart rate or ratios (all correlations *r* < 0.15, *p* > 0.05). Regarding correlations with heart rate, participants showing a stronger increase in heart rate in the arithmetic, compared to the breath focus condition, also showed a more pronounced increase in the occurrence of ratios between 6 and 8.5 (r (19) = 0.75; *p* < .001) and inversely, a more pronounced decrease in ratios between 10 and 13.5 (r (19) = −0.73; *p* < .001) in the arithmetic, compared to the breath focus condition. Correlations between all reported variables are visualized in **Supplementary Figure 3**.

## 4 DISCUSSION

In this study we investigated neural and cardiac electrophysiology in the context of a breath focus period and a stress-inducing arithmetic task in 21 young adults, who had no prior experience with meditation practices. Besides evaluating neural and cardiac domains individually, we analyzed the relationship between both signals across conditions. To that end, we analyzed cross-frequency relationships between the alpha oscillations (8-14 Hz) and heart rate (0.65-2 Hz). Overall, significant differences between conditions were evident both in the cardiac and neural systems, as well as in terms of their relationship.

Regarding condition-related changes in the cardiac domain, heart rate was consistently lower during the breath focus task compared to the arithmetic task. These results are in line with previous literature showing increased heart rate during stressful tasks such as arithmetic calculations (Sun et al., 2019; Watford et al., 2020), but also other tasks including speech stressors (Woody et al., 2017) and the trier social stress test (TSST, Hoge et al., 2018). Several studies have also explored the effects of meditation practices on cardiac physiology, yielding contradictory findings. In a study by Sun et al., (2019), individuals with no experience in mindfulness practices took part in a stress-inducing mental arithmetic task followed by a breathing space meditation task, which resulted in a significant decrease in mean heart rate. Similarly, other studies have found reductions in heart rate from baseline to meditation practices based on the breath in novices (Bortolla et al., 2022; Ooishi et al., 2021). Lumma and colleagues (2015) reported varying effects on cardiac activity depending on the type of trained and practiced meditation. In the case of breathing meditation, heart rate slightly, but non-significantly, increased from the initial weeks of training to the last, whereas for two other types of meditation (observing thoughts and loving-kindness meditation), there was a significant increase, also significantly higher than for breath focus. In the latter study, assessed experienced effort across mediation practices correlated with changes in heart rate (Lumma et al., 2015). Other studies, however, did not discover differences in heart rate during meditation compared to baseline (Guo et al., 2022; Jiang et al., 2020), in Tibetan Buddhist monk practitioners with extensive meditation experience, or between monks and controls during rest (Jiang et al., 2020). In terms of relaxation effects, Balban et al., (2023) recently compared a typical breath focus meditation practice with specific breathwork exercises (i.e., cyclic sighing, box breathing, cyclic hyperventilation with retention) aimed at relaxation. Whereas a reduction in heart rate was found consistently across exercises, the effects were more pronounced for the exercises aimed at relaxation compared to the meditation practice.

With respect to the neural domain, our results showed faster individual alpha frequencies during the arithmetic task compared to the breath focus condition, at right- and central-frontal locations (Fz, F4, Cz). According to the literature, alpha peak frequency accelerates during tasks involving increasing cognitive effort (Haegens et al., 2014; Mierau et al., 2017), including arithmetic tasks performed by novice (Rodriguez-Larios & Alaerts, 2019) and expert meditators (Rodriguez-Larios, Faber, et al., 2020). Similar to our results, previous studies have found alpha peak frequency to decelerate during meditation practices (Aftanas & Golocheikine, 2002; Irrmischer et al., 2018; Mierau et al., 2017; Saggar et al., 2012), and also compared to an arithmetic task (Rodriguez-Larios, Faber, et al., 2020) and rest in highly experienced meditators (Rodriguez-Larios et al., 2021).

Studies investigating the electrophysiological correlates of different states and tasks such as meditation practices and mental arithmetic, have commonly explored signals in isolation to each other, thereby failing to account for the complex interaction that takes place between the cardiac and neural systems in the human body. Here, we applied the cross-frequency ratio paradigm proposed by Klimesch (2018), whereby a functionally relevant task-specific architecture of human physiological oscillators is proposed. During performance of our arithmetic task, participants displayed a significantly higher occurrence of ratios between 6 and 8.5 across the entire scalp. In contrast, the frequency of occurrence of ratios between 10 and 13.5 was significantly higher during the breath focus period. Hence, our results demonstrate that there are task-specific alpha: heart rate cross-frequency relationship profiles. In particular, the profile found for the arithmetic task includes the integer ratio 8. It is emphasized by Klimesch (2018), that such harmonic cross-frequency relationship between the neural and cardiac domains enables the possibility of phase-coupling between the signals, yielding a functionally relevant arrangement for cognitively demanding tasks. To date, only one other study has examined Klimesch’s framework with respect to neural and visceral rhythms across different tasks, including a memory task, rest and sleep (Rassi et al., 2019). In that study, while alpha, heart rate and breathing frequencies exhibited harmonic cross-frequency relationships during a memory task, non-harmonic relationships were found during sleep. Interestingly, the authors interpret the lack of harmonicity between neural and bodily rhythms during sleep as a lack of body awareness. Regarding non-harmonic interactions between alpha and heart rate, the value proposed to exert maximal decoupling between these oscillators results from multiplying the integer 8 by the irrational number golden mean (1.618…), which yields the value 12.94 (Klimesch, 2018). This value and its multiples have been proposed as the most mathematically effective configuration to avoid spurious phase meetings between pairs of oscillators (Klimesch, 2018; Pletzer et al., 2010). During our breath focus task, participants displayed low percentages of values close to 12.94, but nonetheless, statistically higher percentages of occurrence when compared to the arithmetic task. Considering that for the breath focus task the highest percentages collapse at values clustered around 9, caution needs to be taken when interpreting the significant cluster pertaining to values closer to 12.94. Taking into account that during both the arithmetic and the breath focus task, both integer and non-harmonic values are present to a similar extent, it is challenging to determine whether our measure aids in distinguishing neural-cardiac functional differences between tasks. It is observable however, that for the breath focus task, ratios cluster to higher values than for arithmetic, and at the right edge of the distribution (around the value 13), there are significantly higher percentages of ratios compared to the arithmetic task. Considering that our participants were novices, this could be interpreted as an early stage approximation to the proposed non-harmonicity neural-cardiac profile. It is possible that considering their lack of expertise with meditation practices, the participants found the task cognitively challenging which could yield mixed results. It would therefore be necessary for future studies to investigate the degree of difficulty and effort experienced by participants with such task, as implemented in previous studies (Lumma et al., 2015). Additionally, future studies would benefit from comparisons with a sample of expert meditators, since it is typical for contemplative neuroscientific studies to find differences depending on the degree of expertise with meditation practices of the sample (Berkovich-ohana et al., 2016; Brandmeyer & Delorme, 2018; Rodriguez-Larios et al., 2021; Rodriguez-Larios, Wong, et al., 2020)

Considering that our primary measure of cross-frequency interactions derives from a frequency ratio calculation (dividing alpha by its simultaneous heart rate value), and that significant differences in both heart rate and alpha were observed across conditions, we conducted surrogate data analyses to assess the robustness of the cross-frequency ratio results. When generating new heart rate time series combining data of both conditions, we found that the significant ratio differences between conditions were fairly abolished. These findings demonstrate that an intact heart rate time series is required for yielding the identified condition-related effects in alpha: heart rate cross-frequency ratios. Permutating physiologically observed alpha time series across conditions, on the other hand, did not completely eliminate identified condition-related effects in alpha: heart rate cross-frequency ratios, indicating that the condition-related difference in these ratios relies more heavily on instantaneous changes in heart rate, and to a lesser extent on instantaneous changes in alpha. Also, conducting additional surrogate analyses that completely disregard the physiologically relevant characteristics of the alpha frequency distributions-by randomly selecting alpha peak frequencies (between 8-14 Hz) while keeping the heart rate time series intact-did not fully eliminate condition-related effects observed in alpha: heart rate cross-frequency ratios. This indicates that, while instantaneous changes in alpha frequency might contribute little to the observed condition-related changes in alpha: heart rate ratios, physiologically recorded alpha frequency variations (as opposed to randomly generated) are a prerequisite for condition-related changes in alpha: heart rate relationships to become evident.

Other studies have used alternative analysis approaches to investigate brain-heart interactions across different tasks. For example, Zanetti and colleagues (2019) studied information dynamics during rest, a mild stress-inducing game and a stress inducing arithmetic task. They found an increase in neural-cardiorespiratory connectivity during mental stress as compared to the other tasks, which they interpret as an increase in the network connections enabling regulatory mechanisms necessary during stressful events. In a later study (Pernice et al., 2021) exploring the same dataset with a different analysis approach, multivariate correlations, an increase in brain-heart interactions was consistently found during the mental arithmetic task, localized in frontal electrode sites. Along the same pattern of results, in a recent study, Candia-rivera et al., (2023) investigated bidirectional neural-cardiac interactions during different levels of mental stress and rest. In sum, the authors found that stress modulates the ascending interaction between cardiac activity and the alpha frequency band, as well as with delta, beta and gamma. This collection of studies show an increase in the interaction between the neural and cardiac subsystems under conditions of mental stress, aligning with our finding that, during the stress-inducing arithmetic task, alpha and heart rate display cross-frequency relationships closer to the integer 8, also proposed to facilitate information exchange between the two physiological subsystems (Klimesch, 2018).

A previous study also explored brain-heart interactions via heartbeat evoked potentials (Jiang et al., 2020) during meditation practice in a group of long-term Tibetan Buddhist meditation practitioners and meditation-naïve matching controls. This study found expert meditators to exhibit lower HEP amplitudes during meditation compared to rest, in a cluster comprising right central and frontal sensors, at 340-360 ms after the R-peak. The authors also performed source reconstruction analyses to elucidate the neural origins of the observed changes, and found the bilateral anterior cingulate cortex (ACC) and superior medial frontal gyrus to be involved. Interestingly, resting state heart rate, in both monks and controls, as well as during meditation in monks, did not differ (see Supplementary materials, Jiang et al., 2020). It is also relevant to note that, besides the expertise in meditation, the here compared studies also differ with respect to the studied type of meditation. Whereas Jiang and colleagues (2020) studied practices involving mantra reciting or light focus on a mental object or image, we studied breath focus. Interestingly, while our type of meditation focuses on monitoring bodily signals, the objects of focus are different from visceral signals in the other study. In fact, Jiang and colleagues (2020) interpret their HEP amplitude difference based on the lack of attention to one’s body signals during their meditation condition. Considering the role of ACC, the authors suggest that there is a reduction of internal conflict derived from the lack of attention to potentially distracting internally generated physiological signals (e.g., cardiac activity) during meditation as opposed to rest. Alternatively, instead of reflecting increased heart-brain interactions, recent systematic reviews have also linked increased HEP to modulations of arousal (Coll et al., 2021). For example, fronto-central HEPs around 200-300 ms after the R wave (Coll et al., 2021), as well as HEPs >400 ms after the R peak (Di et al., 2015; Gray et al., 2007; Schulz et al., 2013) have been found to be associated with modulations of arousal.

In order to further investigate the contributions of heart rate and alpha condition-differences, we explored relationships between individual physiology (heart rate and alpha) and cross-frequency ratios. Pearson correlations revealed significant relationships with heart rate differences across conditions. Increases in heart rate from breath focus to the arithmetic task were accompanied by increases in the occurrence of ratios within the cluster associated to higher occurrences during the arithmetic task (ratios between 6.5 and 9). Conversely, the increase in heart rate was associated to decreases in the occurrence of ratios within the cluster associated to the breath focus task (ratios between 10 and 13.5).

Some limitations are worth considering within the current study. Although changes in individual alpha frequency had a specific topographic distribution, our result pattern with the cross-frequency ratio approach yielded very similar results across the entire scalp. Besides the portrayed relevance of changes in heart rate to yield these widespread condition-related differences, the available amount of electrodes in our EEG setup (21) did not reliably allow for a source localization analysis, therefore not enabling a characterization of different brain regions showing differential cross-frequency ratio effects. Future research in this direction would benefit from employing higher density EEG setups or magnetoencephalography to study cross-frequency interactions. Additionally, the here investigated breath focus condition did not allow for separation of breath focus from mind-wandering episodes, thus potentially entailing a mix of two distinct cognitive states within the same condition. However, research with our same participants during an experience sampling task (Rodriguez-Larios & Alaerts, 2020) revealed that mean alpha frequency was significantly higher during mind-wandering compared to breath focus periods. Considering that mind-wandering is proposed to share neural correlates with effortful cognition (Rodriguez-Larios & Alaerts, 2020) like arithmetic tasks (Rodriguez-Larios, Faber, et al., 2020; Rodriguez-Larios & Alaerts, 2019), and that we found a consistent decrease in alpha peak frequency during the breath focus condition, we hypothesize that a potential division between breath focus and mind-wandering periods would have in fact yielded stronger results in the same direction we found here. Lastly, as indicated in previous studies (Jiang et al., 2020), considering that other visceral rhythms such as the gastric rhythm have been shown to account for alpha rhythm changes (Richter et al., 2017), future studies should include a more comprehensive assessment of physiological relationships including gastric activity.

## 5 CONCLUSION

The current study elaborates on the identification of relevant physiological markers during conditions of mental stress and meditation practices. In a pursue to acknowledge the systemic nature of physiological and cognitive processes, we place an emphasis on analyses addressing the interaction between the neural and cardiac subsystems, and find informative profiles depending on the condition. This is, to our knowledge, the first study addressing relationships between neural and cardiac physiology during meditation in novice practitioners. Our results can help elucidate which processes underlie the benefits of meditation practices at early stages of training, as well as can be used to inform novel parameters for the development of neuro- and biofeedback protocols for self-regulation trainings.

## CREDIT STATEMENT

**Javier R. Soriano:** Conceptualization, Methodology, Software, Formal Analysis, Writing-Original Draft, Visualization, Supervision, Project Administration**. Julio Rodriguez-Larios:** Software, Validation, Formal Analysis, Investigation, Resources, Data Curation, Writing-Review & Editing**. Carolina Varon:** Supervision, Writing-Review & Editing. **Nazareth Castellanos**: Supervision, Writing-Review & Editing**. Kaat Alaerts:** Conceptualization, Writing-Review & Editing, Supervision, Funding Acquisition.

## DATA AND CODE AVAILABILITY STATEMENT

Raw EEG and ECG data as well as MATLAB code will be made publicly available in the KU Leuven Research Data Repository upon request and/or after acceptance for publication.

## SUPPLEMENTARY MATERIALS

**Supplementary Figure 1.**
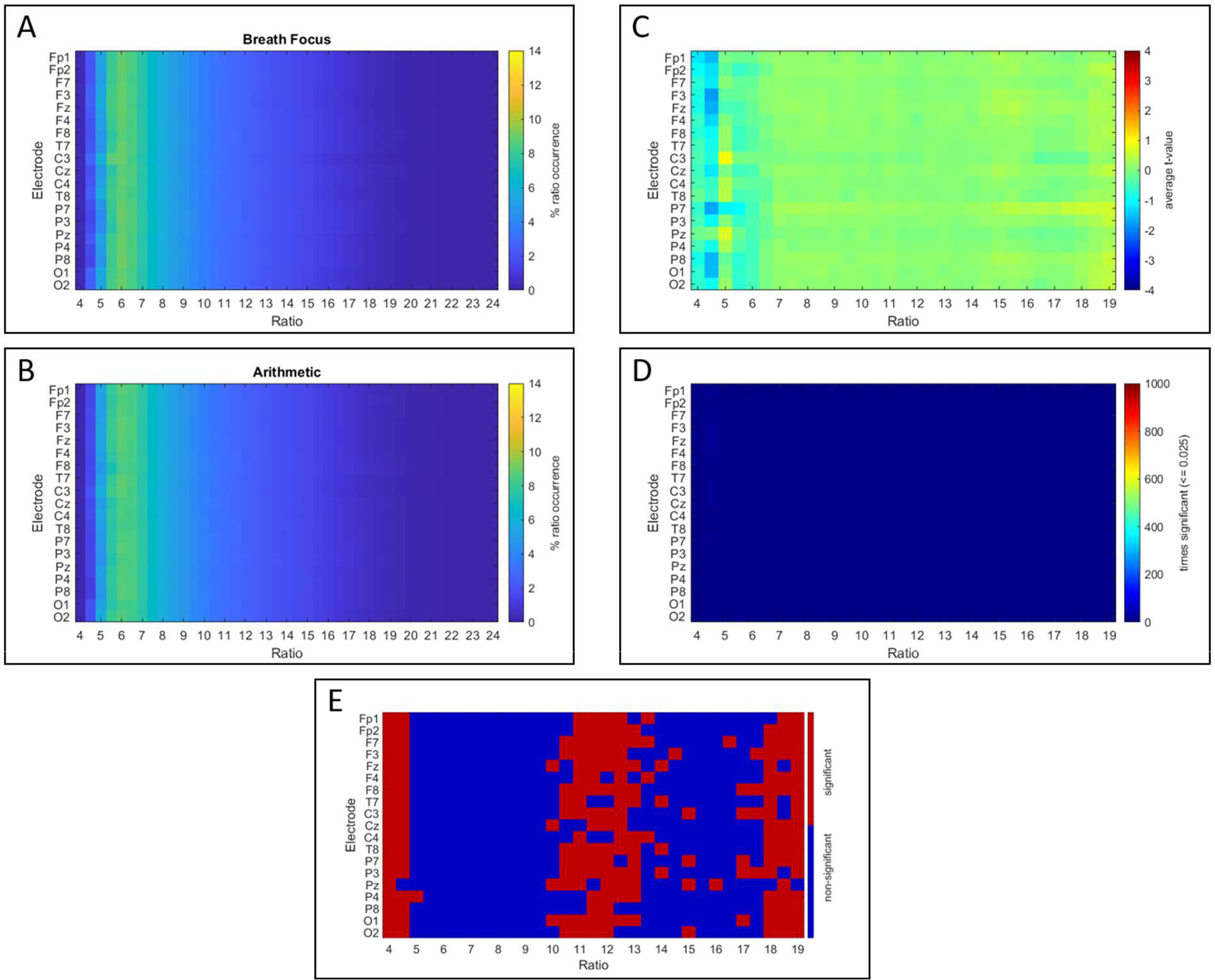
Condition-related differences in alpha: heart rate cross-frequency ratios with randomly generated heart rate data. **A)** visualizes the distribution of the percentage of each ratio occurrence for the breath focus condition, separately for each electrode (y-axis) and cross-frequency ratio (x-axis) (ratios ranging between 4 and 24). **B)** visualizes the distribution of the percentage of each ratio occurrence for the arithmetic condition. Note that the percentage of occurrence of each ratio was calculated separately per participant and electrode, and subsequently averaged across participants. **C)** visualizes the colormap of the average t-values across the 1000 surrogate calculations and permutation tests estimating the condition-related effect (arithmetic versus breath focus condition), separately for each electrode (y-axis) and surrogate cross-frequency ratio (x-axis). Permutation analyses are visualized for ratios ranging between 4 and 19, since the occurrence of ratios below 4.5 or above 18.5 was < 0.01 %. **D)** visualizes the accumulated times that each of the cells (electrode-ratio t-test value) was significant, i.e., over the 1000 surrogate data generations, **E)** shows the significance of the original t-values (plotted in Figure 2D) against the t-value distribution derived from the 1000 surrogate generations.

**Supplementary Figure 2.**
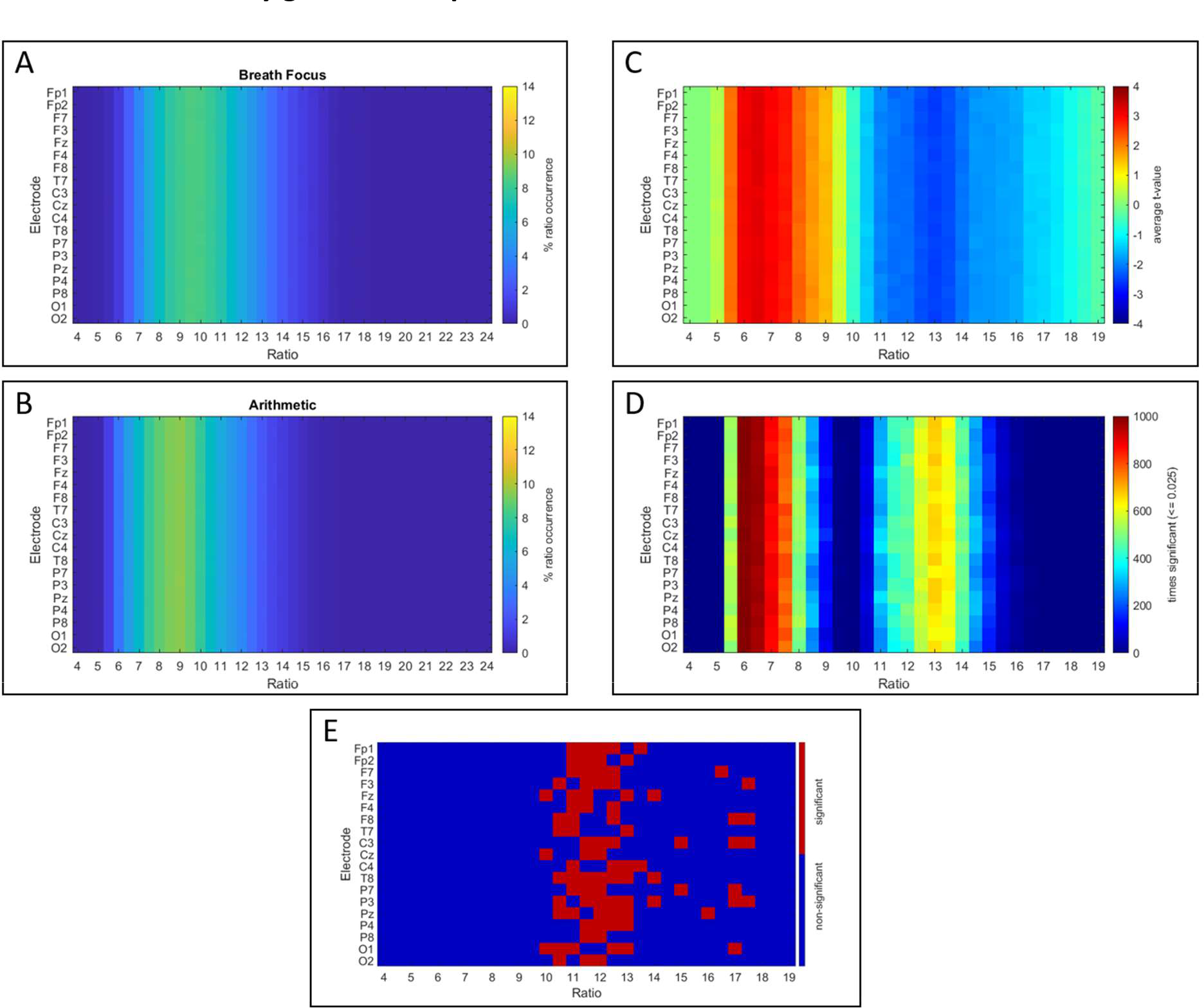
Condition-related differences in alpha: heart rate cross-frequency ratios with randomly generated alpha data. **A)** visualizes the distribution of the percentage of each ratio occurrence for the breath focus condition, separately for each electrode (y-axis) and cross-frequency ratio (x-axis) (ratios ranging between 4 and 24). **B)** visualizes the distribution of the percentage of each ratio occurrence for the arithmetic condition. Note that the percentage of occurrence of each ratio was calculated separately per participant and electrode, and subsequently averaged across participants. **C)** visualizes the colormap of the average t-values across the 1000 surrogate calculations and permutation tests estimating the condition-related effect (arithmetic versus breath focus condition), separately for each electrode (y-axis) and surrogate cross-frequency ratio (x-axis). Permutation analyses are visualized for ratios ranging between 4 and 19, since the occurrence of ratios below 4.5 or above 18.5 was < 0.01 %. **D)** visualizes the accumulated times that each of the cells (electrode-ratio t-test value) was significant, i.e., over the 1000 surrogate data generations, **E)** shows the significance of the original t-values (plotted in Figure 2D) against the t-value distribution derived from the 1000 surrogate generations.

**Supplementary Figure 3.**
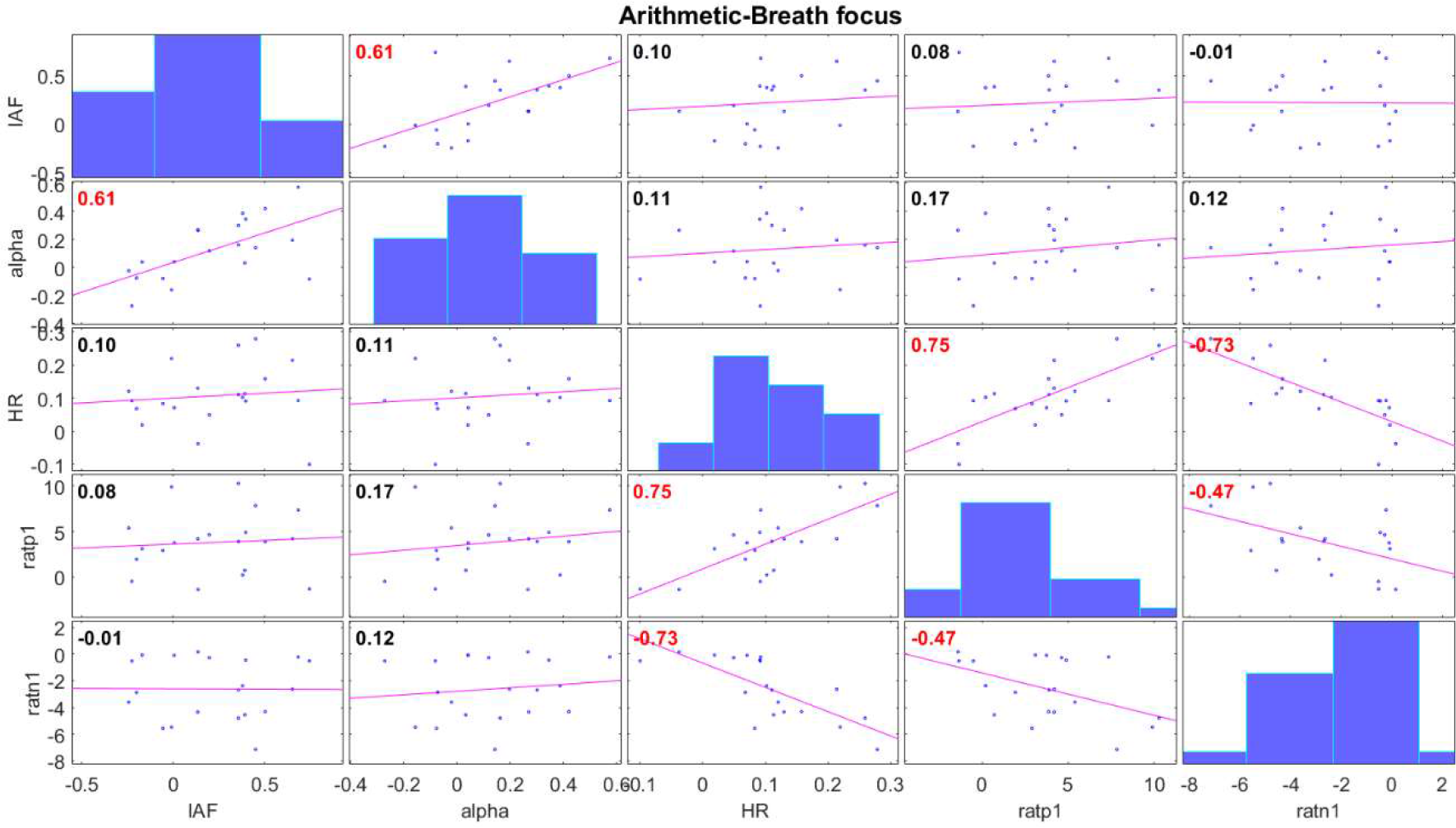
Correlation matrix of average physiological values of the difference arithmetic minus breath focus condition. Following the diagonal downwards, IAF indicates the distribution of average difference values of Individual Alpha Frequency per participant across electrodes, alpha indicates the average difference instantaneous alpha values per participant across electrodes and heart rate the average heart rate instantaneous difference values per participant. Ratp1 indicates the average difference in percentage of ratio occurrence (with respect to all possible alpha: heart rate ratios) for the positive significant cluster of ratios between 6.5 and 9, and Ratn1 the average difference in percentage of ratio occurrence (with respect to all possible alpha: heart rate ratios) for the negative significant cluster of ratios between 10 and 13.5. Red values inside scatterplots indicate significant correlations (p<0.05).

**Supplementary table 1.** Available epochs per condition. Number of Arithmetic (AR raw) and Breath focus (BF raw) available raw epochs, second and third columns respectively. Number of available epochs after correction for arithmetic (AR corrected) is shown in the fourth column and breath focus (BF corrected) in the fifth column. The last column (Final) indicates the total selected epochs for both conditions for analyses.

| Participant | AR raw | BF raw | AR corrected | BF corrected | Final matched |
| --- | --- | --- | --- | --- | --- |
| P1 | 186 | 298 | 171 | 281 | 171 |
| P2 | 238 | 304 | 238 | 291 | 238 |
| P3 | 216 | 302 | 119 | 273 | 119 |
| P4 | 220 | 305 | 196 | 266 | 196 |
| P5 | 289 | 300 | 181 | 205 | 181 |
| P6 | 101 | 312 | 44 | 305 | 44 |
| P7 | 88 | 297 | 86 | 272 | 86 |
| P8 | 385 | 298 | 379 | 297 | 297 |
| P9 | 112 | 299 | 95 | 273 | 95 |
| P10 | 133 | 307 | 122 | 284 | 122 |
| P11 | 195 | 298 | 134 | 190 | 134 |
| P12 | 279 | 320 | 239 | 314 | 239 |
| P13 | 127 | 300 | 120 | 249 | 120 |
| P14 | 234 | 299 | 195 | 243 | 195 |
| P15 | 175 | 318 | 76 | 226 | 76 |
| P16 | 149 | 299 | 72 | 296 | 72 |
| P17 | 199 | 296 | 168 | 223 | 168 |
| P18 | 263 | 299 | 184 | 183 | 184 |
| P19 | 112 | 299 | 110 | 296 | 110 |
| P20 | 119 | 304 | 99 | 302 | 99 |

In **Supplementary Figure 3**, IAF indicates the distribution of difference (from breath focus to arithmetic) values of IAF per participant across electrodes, alpha indicates the average difference instantaneous alpha values per participant across electrodes and heart rate the average heart rate instantaneous difference values per participant. Ratp1 indicates the average difference in percentage of ratio occurrence (with respect to all possible alpha: heart rate ratios) for the positive significant cluster of ratios between 6.5 and 9, and Ratn1 the average difference in percentage of ratio occurrence (with respect to all possible alpha: heart rate ratios) for the negative significant cluster of ratios between 10 and 13.5.

